# Impact of human CD8+ T cell senescence on ^89^Zr radiolabelling and homing properties

**DOI:** 10.1101/2023.02.04.23285413

**Authors:** Jonas Bystrom, Amaia Carrascal-Miniño, George P Keeling, Truc Pham, Conor Garrod- Ketchley, Johannes Schroth, Rafael T. M. De Rosales, Samantha YA Terry, Sian M. Henson

## Abstract

The ability of CD8+ T cells to protect against infection and malignant transformation diminishes with age. Novel means to assess cellular functional deficits *in vivo* are being made available such as total-body positron emission tomography (PET) and radiotracers with long half-lives. Here, we determined radiolabeled human CD8+ T cells isolated from young and old individuals with zirconium-89 (^89^Zr) and assessed their biological status *in vitro* and distribution *in vivo*.

**Methods:** Fresh and cryopreserved CD8+ T cells from young and old individuals were labelled with varying activities of [^89^Zr]Zr-oxine and assessed for DNA damage and survival, subsequent to *in vitro* culture in the presence or absence of antioxidants. ^89^Zr-labelled CD8+ T cells were injected intravenously in NSG mice and whole-body cell migration assessed using PET imaging.

**Results:** Fresh and cryopreserved CD8+ T cells showed no difference in ability to be labelled with ^89^Zr, radionuclide retention, or CD8+ T cell phenotype. ^89^Zr induced partial cell death and DNA damage, which was no longer detectable visible after four days. The level of DNA repair observed in old samples was highly variable. ^89^Zr efflux from cells, seen *in vitro*, did not occur *in vivo*. Longitudinal PET imaging indicated that CD8+ T cells from old individuals accumulated in tissues at a slower rate than those isolated from young individuals.

**Conclusion:** We have established a strategy to label and track the biodistribution of cryopreserved CD8+T cells. Further study is required to understand differences in migratory behaviour of CD8+ T cells isolated from old and young individuals.

## Introduction

Preclinical positron emission tomography (PET) imaging allows for non-invasive total body imaging with a 10- to 40-fold increase in sensitivity compared to conventional systems (1, 2). PET therefore provides the means to analyse immune cell localisation in the whole body in detail, using small numbers of radionuclide-labelled cells (3, 4). Using radionuclides for PET imaging makes the detection of discrete cell populations within defined tissue locations possible (5).

PET radionuclides with a long half-life, such as ^89^Zr (*t*_1/2_ = 78.4 hr), can be used for (pre)clinical imaging and enable the detection of cells longitudinally from repeat PET scans. ^89^Zr-labelling can be achieved either indirectly or directly. ^89^Zr-labelled antibodies have been used to demonstrate the role of CD8+ T cells in tumour immunity and their response to therapy (4, 6). Direct labelling of immune cells provides increased sensitivity and specificity (7, 8) and is achieved through the chelation of ^89^Zr with four oxine (8-hydroxyquinoline) molecules to generate a metastable complex that can cross cell membranes. This complex dissociates inside cells, depositing ^89^Zr bound to intracellular proteins (7, 8). A caveat with direct labelling is that lymphocytes are prone to radiation damage, senescence, and cell death (9, 10). However, CAR T cells labelled with ^89^Zr showed no reduction in cytokine production, migration, tumour cytotoxicity, or *in vivo* antitumor activity when subjected to 70 kBq/10^6^ cells (11).

CD8+ T cells can be subdivided into different populations varying in their functional and homing preferences (12). Central memory (CM) CD8+ T cells were shown to provide longer-lasting protective immunity than the effector memory (EM) subset (13). Older individuals, who are more susceptible to malignancies, have an increased proportion of the EM CD45RA re-expressing (EMRA) CD8+ T cells, which display reduced proliferative capacity, higher levels of DNA damage and an altered migratory potential (14). It remains unexplored whether this EMRA subpopulation responds differently to cell labelling and whether homing abilities are altered. Additionally, further characterization of these EMRA T cells will be essential if ^89^Zr is to be used clinically, specifically, whether cryopreservation influences CD8+ T cell phenotype, together with their ability to be labelled and retain ^89^Zr. This is important as most chronic diseases requiring analysis by PET generally occur at an older age (6). Therefore, it is of interest to determine whether the build-up of EMRA T cells with age influences the ability of ^89^Zr labelling for imaging.

Here, we determined radiolabeled human CD8+ T cells isolated from young and old individuals with [^89^Zr]Zr-oxine and assessed their biological status *in vitro* and distribution *in vivo*.

We show in this study that cryopreservation does not influence the ability of CD8+ T cells to be labelled with ^89^Zr. Cryopreserved ^89^Zr-CD8+ T cells were readily detected using PET imaging and tracked for three days in mice. However, ^89^Zr retention in CD8+ T cells from old individuals was lower than cells taken from young people. All CD8+ T cells experienced initial DNA damage which was repaired during *in vitro* culture. However, DNA damage remained high in CD8+ T cells from older individuals. Labelling of CD8+ T cells and longitudinal assessment by *in vivo* PET scanning indicated retention of ^89^Zr but different migratory patterns when using young and old T cells. This study suggests that the age of CD8+ T cells should be taken in account during longitudinal PET studies as it may influence the DNA-damage response to ^89^Zr, and thereby the migratory capacity of the studied cells.

## Materials and Methods

### Reagents and animals

[^89^Zr]Zr(oxalate) was purchased from Perkin Elmer. Ficoll Paque was purchased from GE Healthcare. PBS, RPMI and ascorbic acid and 2-mercaptoethanol (2-ME) was purchased from Merck Millipore. IL-2 and anti-CD3 (OKT3) was purchased from BioLegend. Anti-CD8 magnetic beads were purchased from Miltenyi. Female Nod *scid* gamma (NSG, NOD.Cg-*Prkdc* ^scid^ *IL2rg^tm1Wjl^*/SzJ) mice aged 6 weeks were purchased from Charles River (UK).

### Ethical Approval

Our clinical protocol was approved by the West London & GTAC Research Ethics Committee (20/PR/0921) and all subjects provided written informed consent. We recruited healthy volunteers in two age ranges, young: 23-58 years of age (n = 8), and old: 69-82 years of age (n = 4). Animal experiments were approved by the UK Home Office under The Animals (Scientific Procedures) Act (1986), PPL reference 7008879 (Protocol 6), with local approval from King’s College London Animal Welfare and Ethics Review Body.

### Cell isolation

Blood (30-60 mL) was obtained from heathy young or old individuals. Peripheral blood mononucleated cells (PBMCs) were isolated using Ficoll Paque. Cells were resuspended in foetal bovine serum (FBS) with 10% dimethylsulfoxide (DMSO) and stored in liquid nitrogen or used directly. Cryopreserved PBMCs were thawed, washed once using RPMI, resuspended in RPMI culture medium and stored at 37°C 1 hr prior to the labelling procedure. Cell number was determined using a haemocytometer prior to cell labelling. CD8+ T cells were isolated from PBMCs using Miltenyi magnetic beads and magnetic column as has been described before (15). CD8+ T cells from cryopreserved samples were isolated upon thawing and then treated as the cryopreserved PBMCs prior to labelling.

### Cell culture

For radiotracer retention and cell proliferation studies, 1 x 10^6^ radiolabelled (or vehicle-treated) PBMCs or CD8+ T cells were cultured for up to 4 days using RPMI, supplemented with 10% FBS, L-Glutamine (2 mM), penicillin and streptomycin (Sigma) and 1 ng/mL IL-2 (Miltenyi). For CD8+ T cell cultures, tissue culture plates were precoated with 0.5 µg/mL anti-CD3 prior to addition of the cells.

### [^89^Zr]Zr-oxine synthesis, cell labelling, and viability

^89^Zr was chelated with oxine forming [^89^Zr]Zr(oxinate)_4_ as previously described (7).

White blood cells (PBMC or separated CD8+ T cells) were washed with PBS (Ca^2+^/Mg^2+^ free), resuspended in PBS and mixed with [^89^Zr]Zr(oxinate)_4_ with a volume ratio not lower than 30:1 (cells: [^89^Zr]Zr(oxinate)_4_), using 3-20×10^6^ cells and 5-100 mBq/cell in various experiments. In some experiments, PBMCs were resuspended in PBS containing 0.1 or 10 mg/mL ascorbic acid or 50 µM 2-mercaptoethanol. The labelling reaction was stored at 37°C or 4°C for 30 min, with agitation every 5 min. After radiolabeling, cells were washed using 50 mL PBS and quantified using a haemocytometer. Radioactivity in the pellets and supernatants of centrifuged and washed cells was then counted using a Wallac Wizard gamma counter to determine cell-associated radioactivity. The percentage of initial activity (%IA) present in cells was then calculated.

The number of viable cells remaining in the culture was determined at 24 and 96 hours using a haemocytometer and trypan blue.

### Flow cytometry

Flow cytometric analysis was used to determine CD8+ T cell phenotype, DNA damage, and apoptosis using the following antibodies: anti-CD8 PerCP (SK1), anti-CD45RA APC (HI1000), anti-CCR7 PECy7 (G043H7), and CD27 FITC (O323) from BioLegend. γH2AX (DNA damage, BioLegend) was used for intracellular staining with FOXP3 staining buffer (BioLegend). Exact CD8+ T cells numbers and subpopulations with and without γH2AX was determined by relating proportions determined by FACS: number of PBMCs in culture to the initial number of PBMCs after the cell labelling reaction. Annexin V-FITC (apoptosis) was used with binding buffer from BioLegend.

All samples were analysed using a FACS Melody (BD Biosciences) and the resulting data examined using FlowJo software (BD Biosciences).

### *In vivo* PET imaging of CD8+ T cells

^89^Zr-labeled CD8+ T cells were injected intravenously into NOD scid gamma (NSG) mice (3×10^6^ cells/animal in 100 μL PBS, 36.9 ± 42.8 kBq, single CD8+ T donor per experiment) under anaesthesia (1.5-2.5% isoflurane in oxygen). Three hours post-injection, mice were re-anaesthetised and placed in a preclinical nanoPET/CT scanner (Mediso) where anaesthesia was maintained and the bed was heated to maintain a normal body temperature. Two hours of PET acquisition (1:5 coincidence mode; 5 ns coincidence time window) were followed by CT. PET-CT was repeated at t = 24 and 72 hr (2 and 3 hr, respectively). Dynamic PET/CT images were reconstructed using a Monte Carlo-based full-3D iterative algorithm (Tera-Tomo, 400-600 keV energy window, 1-3 coincidence mode, 4 iterations and subsets) at a voxel size of (0.4 × 0.4 × 0.4) mm^3^ and corrected for attenuation, scatter, and decay. Images were co-registered and analysed using VivoQuant v.3.0 (InVicro LLC) capturing maximum intensity projection (MIP) and transverse plane images. If required, background, not associated with anatomy, were manually removed. Regions of interest (ROIs) were drawn covering lung, liver and spleen structures based on CT image data and activity accumulation determined.

### *Ex vivo* biodistribution

Mice from imaging studies were used for biodistribution studies at 72 hours post injection. After culling, organs were dissected, weighed, and γ-counted together with standards prepared from a sample of injected material. The percentage of injected activity per gram injected dose per gram (%IA/g) of tissue was calculated.

### Statistical analysis

Statistical analysis was performed using GraphPad Prism version 9.5. A Mann-Whitney U test was used to compare differences between groups and Wilcoxon signed rank test within groups, as data was not normally distributed. ANOVA followed by Tukey post hoc testing was used when comparing more than two groups. Data was expressed as mean ± SD and statistically significant differences represented using the following notation: * = p < 0.05, ** = p < 0.01, *** = p < 0.005 and **** = p < 0.001.

## Results

A previously established kit-formulation was used to create [^89^Zr]Zr(oxinate)_4_ with a radiochemical yield of 82.3 ± 7.7 % (Figure 1A) (7). We found no difference in the labelling efficiency of freshly isolated PBMCs compared to bead-purified CD8+ T cells (Figure 1B). Therefore, PBMCs rather than isolated CD8+ T cells were used for optimisation and flow cytometry-based experiments. Additionally, cryopreservation did not influence the radiolabelling efficiency of young PBMCs (Figure 1C). Cryopreserved PBMCs from young and old participants were then labelled with [^89^Zr]Zr(oxinate)_4_ and despite showing more than 50% loss of cells during the 30-minute labelling process (data not shown), no difference in radiolabelling was observed between PBMCs taken from young and old individuals (Figure 1D). Therefore, we concluded that neither cryopreservation nor age influenced cell labelling efficiency. However, differences between young and old cryopreserved PBMCs did become apparent when the cells were cultured. Cryopreserved PBMCs from young participants retained significantly more of the initial activity than cells isolated from older participants, assessed at days one and four post cell labelling (Figure 1E). Overall, the data shows no age-related effect on the ability of ^89^Zr to radiolabel immune cells, although PBMCs from older volunteers retained less radionuclide during *in vitro* culture.

**Figure 1.**
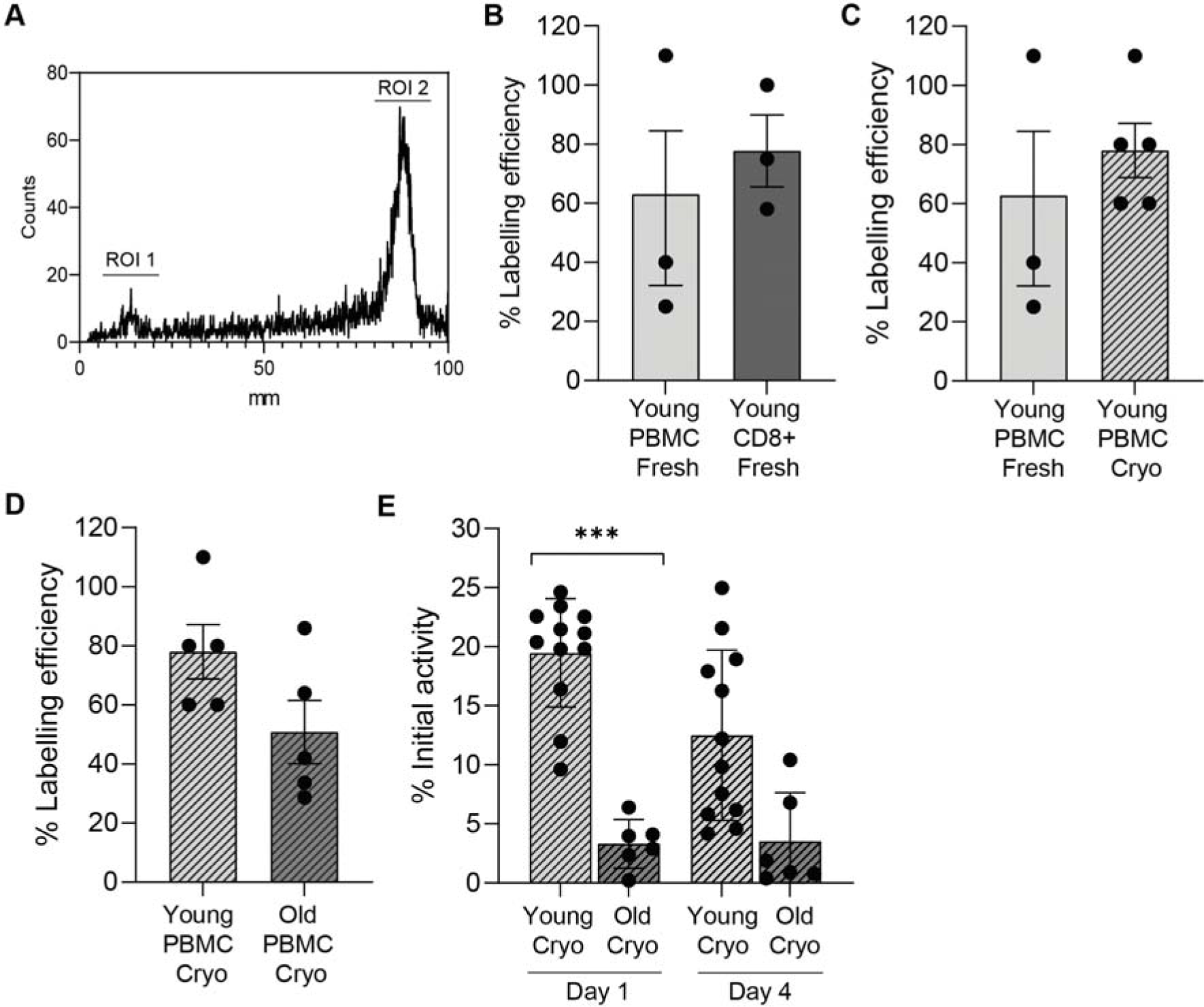
**A.** Example of radio-TLC to determine [^89^Zr]Zr-(oxinate)_4_ chelation efficiency, with region of interest (ROI) region 1 being free ^89^Zr (5.9%) and ROI 2 being [^89^Zr]Zr-(oxinate)_4_, (94.1%). [^89^Zr]Zr-(oxinate)_4_ cell labelling efficiency with young fresh PBMCs and CD8+ bead-isolated T cells (n = 3), **B**; with young fresh (n = 3) and cryopreserved PBMCs (n = 5), **C**; and young cryopreserved PBMCs (n = 5) and old cryopreserved PBMCs (n = 5), **D.** Percentage ^89^Zr remaining associated with cells based on initial activity (%IA) following cell culture at day one and four in young cryopreserved PBMCs (n=12) and old cryopreserved PBMCs (n=6), **E.**

We then assessed how radiolabelling PBMCs isolated from young and old individuals with ^89^Zr influenced their survival and DNA damage response. Irrespective of whether PBMCs were isolated from young or old individuals all ^89^Zr-labelled PBMCs behaved equally; there was a sharp decline in the number of CD8+ T cells at day 1 post radiolabelling after which cell numbers then remained stable for the duration of culture (Figure 2A). We also followed CD8+ T cell numbers post ^89^Zr labelling and observed a similar trend (Figure 2B). The number of CD8+ T cells from old individuals was lower than that of young but was not altered during culture. This decline in CD8+ T cells with age has been reported before and is likely not related to the radiolabelling itself (16). We further explored this immune cell loss using Annexin V to determine the amount of cell death following labelling. The addition of ^89^Zr resulted in a significantly higher proportion of CD8+ Annexin V+ T cells than untreated cells, with the amount of apoptotic CD8+ T cells increasing further after 4 days in culture (Figure 2C).

**Figure 2.**
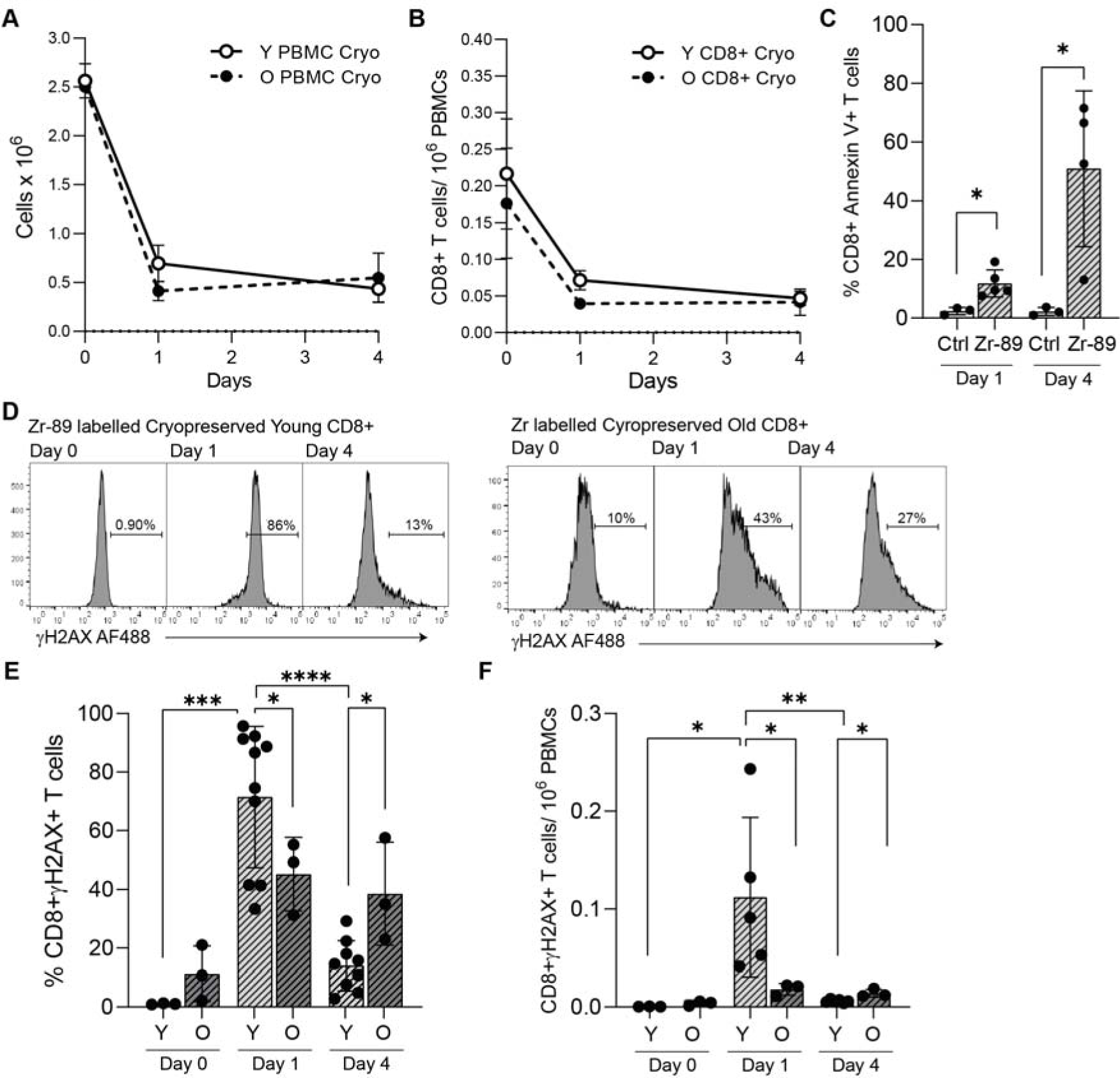
Enumeration of cryopreserved PBMCs isolated from young or old individuals following ^89^Zr labelling and at one- and four-days post radiolabelling (n= 3). **B.** Number of ^89^Zr labelled CD8+ T cells per 10^6^ PBMCs in culture after one and four days. **C.** Proportion (%) of Annexin V+ cells among control and ^89^Zr labelled CD8+ T cells identified in PBMCs isolated from young people, at one and four days of culture (n = 3-5). **D.** Flow histograms showing γH2AX expression gated on CD8+ T cells among ^89^Zr labelled PBMCs taken from young and old people at one and four days of culture. **E.** Bar-graphs of γH2AX in CD8+ T cells following ^89^Zr labelling and culture (n = 3). **F.** γH2AX-positive CD8+ T cells normalised per 10^6^ PBMCs following ^89^Zr labelling and culture (n = 3-5).

To determine whether intracellular ^89^Zr altered the DNA damage response in CD8+ T cells from young and old individuals, an assessment of the double strand break (DSB) associated histone γH2AX was made (Figure 2D). At baseline, ^89^Zr-labelled CD8+ T cells from old individuals tended to have more *ex vivo* DNA damage than young individuals (Figure 2E). After one day of culture, the presence of the ^89^Zr label was found to induce high levels of DNA damage to both young (71.6 ± 24.1% γH2AX+) and old (45.2 ± 12.5% γH2AX+) CD8+ T cells, with significantly more damage being generated in the CD8+ T cells from young people (Figure 2E). At day four, the proportion of young γH2AX+ CD8+ T cells had significantly declined. However, for old labelled CD8+ T cells the γH2AX-level remained high (p = 0.018, Figure 2E). We then adjusted the data to consider the starting amount of PBMCs that underwent ^89^Zr labelling, and also the change in CD8+ T cell subset composition. We confirmed that γH2AX+ in young ^89^Zr labelled CD8+ T cells in cell culture increased at day one but returned to a pre-label level by day 4 (p = 0.016, Figure 2F). As the CD8+ T cells from old individuals were less abundant following the labelling procedure when enumerated they failed to show the same increase in DNA damage as young cells. However, the number of γH2AX+ CD8+ T cells did not decline, failed to repair and persisted in culture (p = 0.024, Figure 2F).

CD8+ T cells are heterogenous and can be subdivided into 4 populations using the cell surface markers CD45RA and CCR7, thereby identifying CD45RA+CCR7+ Naïve, CD45RA-CCR7+ central memory (CM), CD45RA-CCR7-effector memory (EM) and CD45RA+CCR7-EMRA T cells. The EMRA population displays senescent-like characteristics and are expanded in old individuals (14). We assessed whether CD8+ T cell subset composition differed following ^89^Zr-labelling (Figure 3A). Although CD8+ EMRA T cells were more numerous in older individuals (Figure 3A, B) the relative subset distribution was not affected by ^89^Zr-labelling (Figure 3B). These results suggest that ^89^Zr-labelling did not influence CD8+ T cell differentiation.

**Figure 3.**
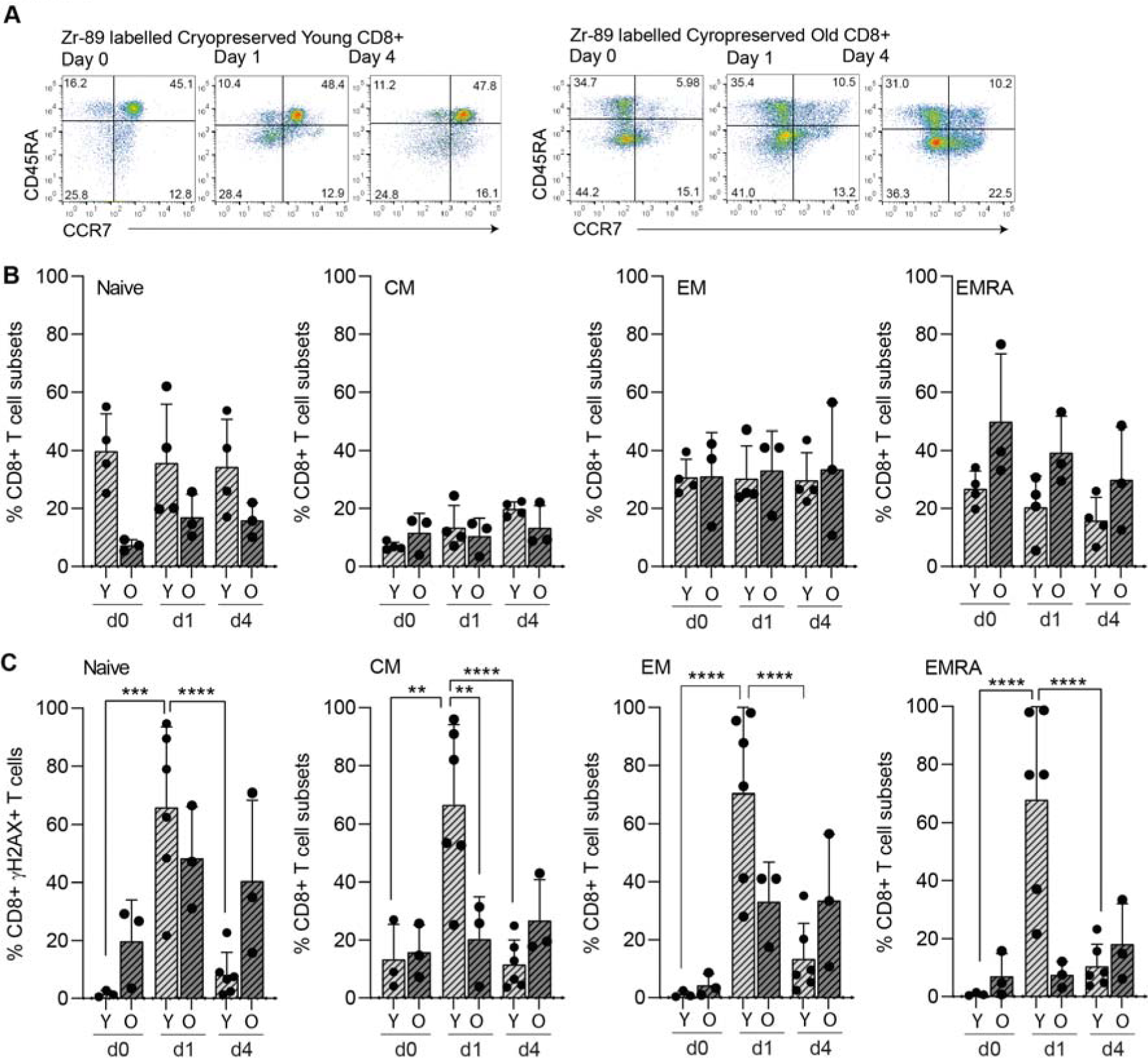
**A.** Representative flow cytometry plots showing CD45RA/CCR7-defined CD8+ T cell subsets before radiolabelling and among radiolabelled young and old T cells at days one and four of cell culture. Naïve: CD45RA+CCR7+, central memory (CM): CD45RA-CCR7+, effector memory (EM): CD45RA-CCR7-, EMRA: CD45RA+CCR7-. **B.** The proportions of each of the CD8+ T cell subsets, (n=3-4). **C.** The DNA damage response in young and old CCR7/CD45RA defined CD8+ T cell subsets. Graphs show the proportion of γH2AX expression in each of the CD8+ T cell subsets, (n=3-6).

We then assessed whether ^89^Zr-induced γH2AX+ cells was overrepresented in any the CD45RA/CCR7 defined CD8+ T cell subsets. While γH2AX+ was found in all four subsets in both young and old individuals, the pattern of DNA damage differed. In young cells, γH2AX was high post-labelling but then declined by day 4. Old cells showed an increase in DSB, which remained high throughout culture (Figure 3C); this could be attributed to the naïve CD8+ T cell population (Figure 3C). In conclusion, DNA damage repair to ^89^Zr in old populations is blunted compared to CD8+ T cells taken from young individuals and is irrespective of subpopulation.

As ^89^Zr labelling leads to enhanced DNA damage and death in CD8+ T cells, we tested ways to prevent this radiation-induced damage. Inhibition of reactive oxygen species has been used previously to prevent damage (17), as has lowering the temperature during the labelling reaction (7). Therefore, 0.1 and 10 mg/mL ascorbic acid and 50 µM 2-mercaptoethanol was included during the [^89^Zr]Zr(oxinate)_4_ labelling reaction. The labelling reaction was also carried out at 4°C. Labelled CD8+ T cells were subsequently kept in culture for analysis of Annexin V binding and expression of γH2AX. A high dose of ascorbic acid (10 mg/mL) and 4°C were found to inhibit the initial [^89^Zr]Zr(oxinate)_4_ uptake (Figure 4A). None of the treatments improved ^89^Zr retention compared to the original conditions performed at 37°C (Figure 4B). Likewise, annexin V and γH2AX expression showed that these strategies were unable to prevent the radiation-induced damage reduced to CD8+ T cells (Figure 4C, D). The phenotype of CD8+ T cells was also assessed with no evidence of changes in the distribution of the 4 subsets in response to any of the radiation damage-prevention methods used (data not shown). In conclusion this data suggested that conventional treatments to scavenge free radicals do not work on CD8+ T cells without simultaneously affecting radiolabelling efficiency..

**Figure 4.**
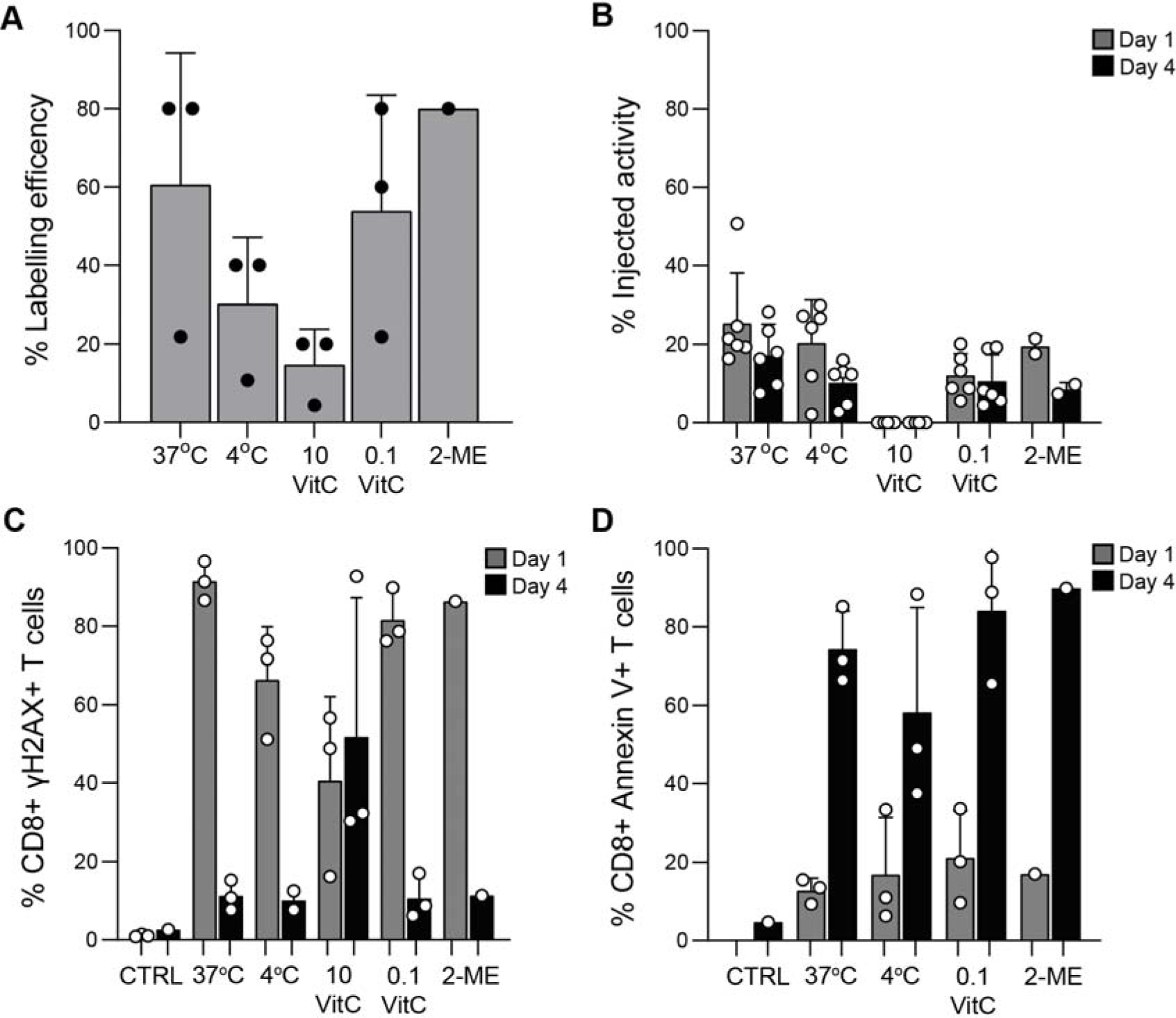
During the [^89^Zr]Zr-(oxinate)_4_ labelling reaction, PBMCs underwent standard incubation (37°C), incubation at 4°C, with 10 or 0.1 mg/mL ascorbic acid at 37°C. or 50 µM 2-ME at 37°C. **A.** Labelling efficiency with [^89^Zr]Zr-(oxinate)_4_ of PBMCs labelled at 50 mBq/cell,. **B.** ^89^Zr retention at one and four days of cell culture. **C.** Annexin V and **D.** γH2AX expression in CD8+ T cells at one and four days of cell culture after ^89^Zr cell labelling (n=3).

To determine whether cryopreservation and age would influence PET imaging studies, 3 × 10^6^ CD8+ T cells from young and old individuals were labelled with [^89^Zr]Zr-(oxinate)_4_ and injected into NOD scid gamma (NSG) mice at a final activity of 67.2 kBq and 6.69 kBq for young and old respectively. CD8+ T cells initially localized to lung and liver (3 hr, Figure 5A-D). While young and old CD8+ T cells gradually accumulated in the spleen, a high proportion of old CD8+ T cells remained in the liver and lung (Figure 5C, D). *Ex vivo* γ-counting at 72 hrs showed the highest concentration of ^89^Zr labelled CD8+ T cells were found in the spleen (Figure 5E). However, CD8+ T cells from old individuals accumulated in the spleen at a significantly lower rate compared to cells isolated from young individuals (Figure 5E). Uptake in other organs showed no major differences between the two groups. Overall, our data indicates an age-related change in the migratory potential of CD8+ T cells from old individuals to lymphoid organs.

**Figure 5.**
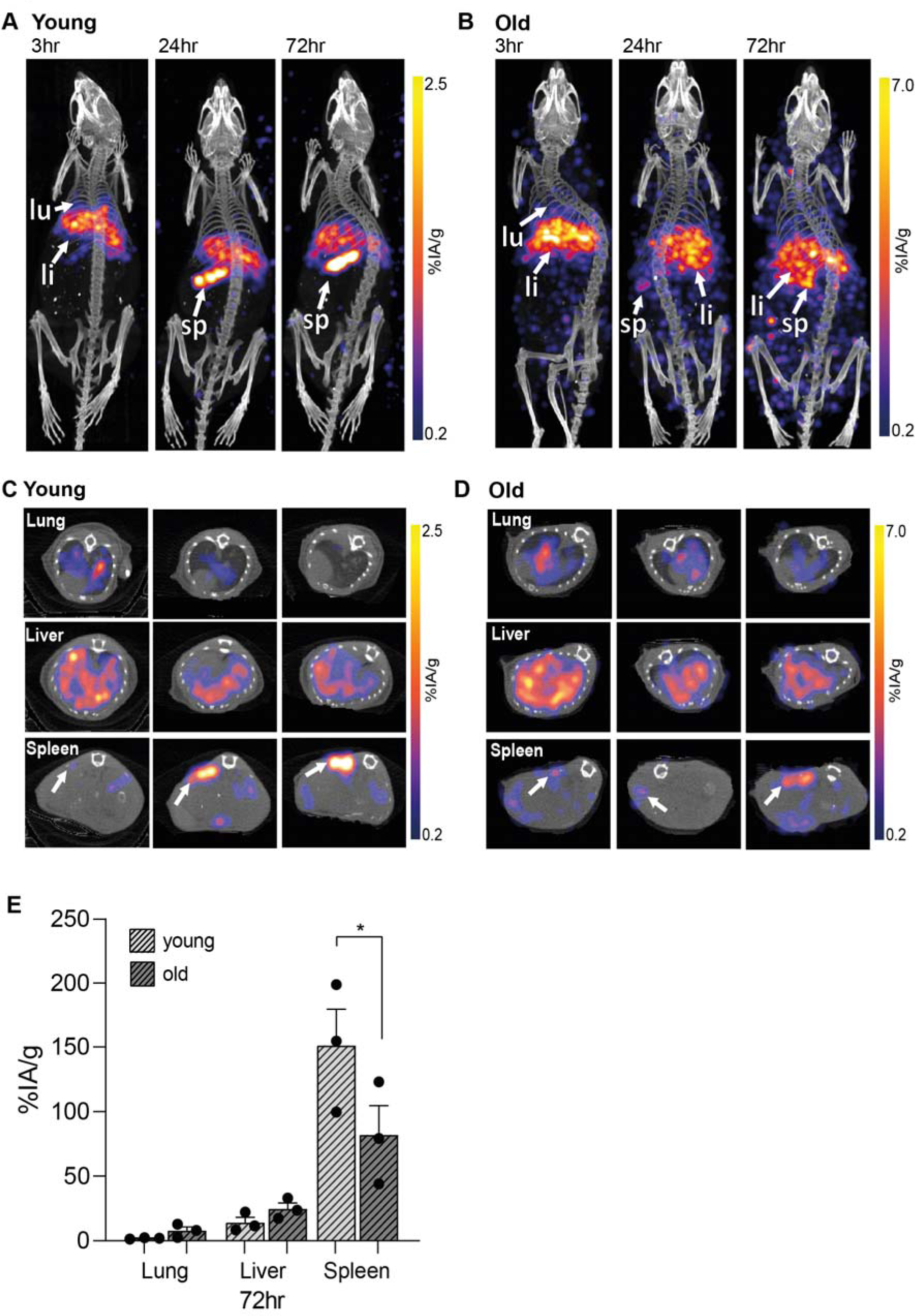
Representative images of NSG mice infused with ^89^Zr-labelled 3 × 10^6^ CD8+ T cells from a young (**A**) an old individual (**B**) at 36.9 ± 42.8 kBq/mouse. Mice were subjected to whole-body preclinical PET at 3 hr (2 hr scan), 24 hr (2 hr scan) and 72 hr (3 hr scan). Transverse sections at time-points 3, 24 and 72hr highlighting regions with lungs, livers and spleens using ^89^Zr labelled cells from young (**C**) and old individuals (**D**). White arrows indicate lung (lu), liver (li) and spleen (sp). Data was reconstructed and analysed using Vivoquant. **E.** *Ex vivo* biodistribution of ^89^Zr-labelled CD8+ T cells isolated from young and old individuals 72 hours after CD8+ T cell administration.

## Discussion

In this study, we have gained important insights that will help better track immune cell subpopulations using the total-body PET modality. Cryopreserved CD8+ T cells, stored in liquid nitrogen, can be labelled with similar efficiency to freshly isolated cells. Cryopreserved ^89^Zr-labelled CD8+ T cells were readily detected using PET imaging and tracked for three days post-infusion. In response to labelling, CD8+ T cells experienced initial DNA damage which was associated with cell death. Furthermore, while a significant proportion of the CD8+ T cells from young individuals underwent DNA repair this was not the case when labelling aged cells. Tissue distribution of CD8+ T cells from young and old individuals suggested that there was an age-dependent difference in organ homing with an alteration in migration dynamics between young and old CD8+ T cells. Hence the old CD8+ T cells remained to a larger extent in the liver, while young cells more efficiently infiltrated the spleen of the mice. Although our *in vitro* studies indicate that CD8+ T cells from old individuals do not retain ^89^Zr as well as the cells isolated from young individuals, the *in vivo* experiments suggest protection *in situ*.

The advent of total body PET with the ability to detect cells with a high sensitivity provides the opportunity to study specific cell subsets that have different homing preferences (2, 18). The high specificity of directly labelled cells have an advantage in that sense, compared to radiolabelled antibodies that can be associated with background (8, 18). In our study, we assessed whether ^89^Zr labelling influenced CD45RA/CCR7 defined CD8+ T cell subsets differently. Although γH2AX was dramatically upregulated in half to more than two thirds of all the cells, the different subset-proportions were likely not altered by the labelling. We also assessed the influence of ^89^Zr labelling on aged CD8+ T cells. This is important as old individuals are more likely than young to undergo PET scan assessment for tracking of immune cells (6). Old individuals have proportionally more EMRAs than young (Figure 3B), which has been well reported previously (16, 19). EMRA T cells display hallmarks of senescence in that they express high levels of KLRG1, dysfunctional mitochondria, increased ROS production leading to more DNA damage and a reduced proliferative capacity (15). The DNA damage response (DDR) to ^89^Zr dose-exposure differed between CD8+ T cells isolated from young and old individuals. Aged naïve T cells had more DNA damage at baseline and went on to upregulate more γH2AX in response to ^89^Zr, which remained present in the cells at day four of culture suggesting an inefficient DNA repair process. However, it has been documented that naïve CD8+ T cells from old individuals are not truly naïve, some originating from the EMRA subset, potentially explaining our observations (20, 21). There are two main pathways to repair DNA double strand breaks (DSB), non-homologous end joining (NHEJ) and homologous recombination (22). Three proteins from the phosphatidylinositol 3-kinase related kinase family are activated by DSBs, ataxia telangiectasia mutated kinase (ATM), ATM-related kinase, and DNA-dependent protein kinase. Early in the DDR, the phosphorylation of histone H2AX (γH2AX) signals the presence of DSB to repair proteins, which in turn aggregate in ionising radiation-induced foci. The DDR is induced by reactive oxygen species and possibly by unprotected DNA during telomere attrition in EMRAs (15, 23). The EMRA did not upregulate γH2AX and could therefore be more prone to cell death.

Unfortunately, our attempts to prevent radiation-induced DNA damage, both using the ROS scavenger ascorbic acids and by undertaking labelling at low temperature, without affective radiolabelling efficacy, were unsuccessful. Our *in vitro* studies suggest that the different *in vivo* migratory preference observed between young and old T cells, may only partly be due to radiation induced changes to CD8+ T cell subsets. DNA damage not only kills older EMRA T cells but could contribute to the localization preference shown here. Indeed, ATM activation has been shown to regulate IL-8 to sustain cell migration (24). Furthermore, the chemokine receptor CXCR2 was found to be upregulated in response to p53 (25). Given that we have shown CXCR2 to be upregulated on CD8+ EMRA T cells from people with type 2 diabetes (26), a condition that induces premature immune senescence, it is not unreasonable to speculate that DNA damage could influence the migratory capability of T cells. Interestingly, in cancer patients, the variability to radiation-induced DDR is different in the CD8+ T cell subsets and has been associated with treatment response. For example, a study of advanced or recurrent hypermutated or microsatellite instability-high, mismatch repair deficient uterine cancer patients showed subset differences in their DDR following check-point blockade, with the EMRA subset showing the highest expression of γH2AX in response to irradiation (27). Furthermore, following checkpoint blockade activation of the ATM downstream of γH2AX was only seen in therapeutic responders. It is likely that such a varied response seen when comparing young and old T cells, in our study, would not only make specific cell populations more prone to radiation-induced death, but potentially also altered cellular function influence homing preferences.

## Conclusion

Cryopreservation did not affect the ^89^Zr labelling potential of CD8+ T cells. However, the DNA damage response following radiolabelling was enhanced in CD8+ T cells isolated from old individuals. Analysis of tissue distribution using ^89^Zr labelled CD8+ T cells from young and old suggest that there is a difference in splenic migration. These findings will have implications for human CD8+ T cell tracking and detailed localization preference using total body PET in response to immune-regulating treatments.

## Data Availability

All data produced in the present study are available upon reasonable request to the authors

## Acknowledgements

We would like to thank Jana Kim and Hannah Greenwood for their help with intravenous injections. This research was supported by Diabetes UK 19/0006057, Barts and the London Charity MGU0536, Royal College of Anaesthetists WRO-2018-0065. Additionally, this work was funded in part by the EPSRC programme for next generation molecular imaging and therapy with radionuclides [EP/S032789/1], the Wellcome/EPSRC Centre for Medical Engineering at King’s College London [WT 203148/Z/16/Z], a Wellcome Trust Multiuser Equipment Grant: A multiuser radioanalytical facility for molecular imaging and radionuclide therapy research [212885/Z/18/Z], Research England Confidence in Collaboration scheme, and the MRC Confidence in Concepts scheme. The nanoPET/CT scanner at KCL was funded by an equipment grant from the Wellcome Trust WT 084052/Z/07/Z. For the purpose of open access, the author has applied a CC BY public copyright licence to any Author Accepted Manuscript version arising from this submission. Finally, we thank the research participants for their commitment to this study and their generous donation of blood samples.

## References

1. Vandenberghe S, Moskal P, Karp JS. State of the art in total body PET. EJNMMI Phys. 2020;7(1):35.

2. Cherry SR, Jones T, Karp JS, Qi J, Moses WW, Badawi RD. Total-Body PET: Maximizing Sensitivity to Create New Opportunities for Clinical Research and Patient Care. J Nucl Med. 2018;59(1):3–12.

3. Tavare R, McCracken MN, Zettlitz KA, Salazar FB, Olafsen T, Witte ON, et al. Immuno-PET of Murine T Cell Reconstitution Postadoptive Stem Cell Transplantation Using Anti-CD4 and Anti-CD8 Cys-Diabodies. J Nucl Med. 2015;56(8):1258–64.

4. Pandit-Taskar N, Postow MA, Hellmann MD, Harding JJ, Barker CA, O’Donoghue JA, et al. First-in-Humans Imaging with (89)Zr-Df-IAB22M2C Anti-CD8 Minibody in Patients with Solid Malignancies: Preliminary Pharmacokinetics, Biodistribution, and Lesion Targeting. J Nucl Med. 2020;61(4):512–9.

5. Yoon JK, Park BN, Ryu EK, An YS, Lee SJ. Current Perspectives on (89)Zr-PET Imaging. Int J Mol Sci. 2020;21(12).

6. Farwell MD, Gamache RF, Babazada H, Hellmann MD, Harding JJ, Korn R, et al. CD8-Targeted PET Imaging of Tumor-Infiltrating T Cells in Patients with Cancer: A Phase I First-in-Humans Study of (89)Zr-Df-IAB22M2C, a Radiolabeled Anti-CD8 Minibody. J Nucl Med. 2022;63(5):720–6.

7. Man F, Khan AA, Carrascal-Minino A, Blower PJ, R TMdR. A kit formulation for the preparation of [(89)Zr]Zr(oxinate)4 for PET cell tracking: White blood cell labelling and comparison with [(111)In]In(oxinate)3. Nucl Med Biol. 2020;90–91:31-40.

8. Man F, Lim L, Volpe A, Gabizon A, Shmeeda H, Draper B, et al. In Vivo PET Tracking of (89)Zr-Labeled Vgamma9Vdelta2 T Cells to Mouse Xenograft Breast Tumors Activated with Liposomal Alendronate. Mol Ther. 2019;27(1):219–29.

9. Zhang J, He T, Xue L, Guo H. Senescent T cells: a potential biomarker and target for cancer therapy. EBioMedicine. 2021;68:103409.

10. Falcke SE, Ruhle PF, Deloch L, Fietkau R, Frey B, Gaipl US. Clinically Relevant Radiation Exposure Differentially Impacts Forms of Cell Death in Human Cells of the Innate and Adaptive Immune System. Int J Mol Sci. 2018;19(11).

11. Weist MR, Starr R, Aguilar B, Chea J, Miles JK, Poku E, et al. PET of Adoptively Transferred Chimeric Antigen Receptor T Cells with (89)Zr-Oxine. J Nucl Med. 2018;59(10):1531–7.

12. Nolz JC. Molecular mechanisms of CD8(+) T cell trafficking and localization. Cell Mol Life Sci. 2015;72(13):2461–73.

13. Tantalo DG, Oliver AJ, von Scheidt B, Harrison AJ, Mueller SN, Kershaw MH, et al. Understanding T cell phenotype for the design of effective chimeric antigen receptor T cell therapies. J Immunother Cancer. 2021;9(5).

14. Akbar AN, Henson SM. Are senescence and exhaustion intertwined or unrelated processes that compromise immunity? Nat Rev Immunol. 2011;11(4):289–95.

15. Henson SM, Lanna A, Riddell NE, Franzese O, Macaulay R, Griffiths SJ, et al. p38 signaling inhibits mTORC1-independent autophagy in senescent human CD8(+) T cells. J Clin Invest. 2014;124(9):4004–16.

16. Li M, Yao D, Zeng X, Kasakovski D, Zhang Y, Chen S, et al. Age related human T cell subset evolution and senescence. Immun Ageing. 2019;16:24.

17. Hosokawa Y, Saga R, Monzen S, Terashima S, Tsuruga E. Ascorbic acid does not reduce the anticancer effect of radiotherapy. Biomed Rep. 2017;6(1):103–7.

18. Kurebayashi Y, Choyke PL, Sato N. Imaging of cell-based therapy using (89)Zr-oxine ex vivo cell labeling for positron emission tomography. Nanotheranostics. 2021;5(1):27–35.

19. Akbar AN, Henson SM, Lanna A. Senescence of T Lymphocytes: Implications for Enhancing Human Immunity. Trends Immunol. 2016;37(12):866–76.

20. Callender LA, Carroll EC, Beal RWJ, Chambers ES, Nourshargh S, Akbar AN, et al. Human CD8(+) EMRA T cells display a senescence-associated secretory phenotype regulated by p38 MAPK. Aging Cell. 2018;17(1).

21. Pulko V, Davies JS, Martinez C, Lanteri MC, Busch MP, Diamond MS, et al. Human memory T cells with a naive phenotype accumulate with aging and respond to persistent viruses. Nat Immunol. 2016;17(8):966–75.

22. Galgano A, Barinov A, Vasseur F, de Villartay JP, Rocha B. CD8 Memory Cells Develop Unique DNA Repair Mechanisms Favoring Productive Division. PLoS One. 2015;10(10):e0140849.

23. d’Adda di Fagagna F, Reaper PM, Clay-Farrace L, Fiegler H, Carr P, Von Zglinicki T, et al. A DNA damage checkpoint response in telomere-initiated senescence. Nature. 2003;426(6963):194-8.

24. Chen WT, Ebelt ND, Stracker TH, Xhemalce B, Van Den Berg CL, Miller KM. ATM regulation of IL-8 links oxidative stress to cancer cell migration and invasion. Elife. 2015;4.

25. Guo H, Liu Z, Xu B, Hu H, Wei Z, Liu Q, et al. Chemokine receptor CXCR2 is transactivated by p53 and induces p38-mediated cellular senescence in response to DNA damage. Aging Cell. 2013;12(6):1110–21.

26. Lau EYM, Carroll EC, Callender LA, Hood GA, Berryman V, Pattrick M, et al. Type 2 diabetes is associated with the accumulation of senescent T cells. Clin Exp Immunol. 2019;197(2):205–13.

27. Muroyama Y, Manne S, Wellhausen N, Oldridge DA, Greenplate AR, Chilukuri L, et al. Induction of a CD8 T cell intrinsic DNA damge and repair response is associated with clinical reponse to PD-1 blockade in uterine cancer.

